# Germline rare deleterious variant load alters cancer risk, age of onset and tumor characteristics

**DOI:** 10.1101/2021.07.02.21259537

**Authors:** Myvizhi Esai Selvan, Kenan Onel, Sacha Gnjatic, Robert J. Klein, Zeynep H. Gümüş

## Abstract

**Background:** Inherited genetic variants play an important role in cancer susceptibility. Recent studies show that rare, deleterious variants (RDVs) in a few genes are critical determinants of heritable cancer risk. Better understanding of germline RDVs will contribute to improved precision prevention, screening and treatment.

**Methods:** We have performed the largest to date jointly processed germline pan-cancer case-control association study from whole-exome sequencing (WES) data of 20,789 participants, split into discovery and validation cohorts. We focused on high-penetrance RDVs based on ClinVar database. To increase the statistical power, we pursued a collapsing approach and compared the cumulative RDV burden at gene and gene-set levels using penalized logistic regression. Next, we investigated how the accumulation of RDVs in an individual (RDV load) is associated with cancer risk. Finally, we studied how personal RDV load in specific gene-sets affected i) age of diagnosis; ii) tumor immune microenvironment; and iii) tumor mutational burden using Mann-Whitney U test and Kruskal-Wallis test.

**Results:** Our results confirm known associations between cancer risk and germline RDVs in *BRCA1/2* genes, and show associations with risk for RDVs in *Fanconi Anemia (FA)*, DNA damage repair (DDR), cancer predisposition (CPD) and somatic cancer driver gene-sets in two independent cohorts. Furthermore, increased personal germline RDV load in these gene-sets increased cancer risk, and once cancer developed, tumor characteristics. Notably, we show that the personal RDV load of an individual in FA, DDR or CPD genes is a potential marker for younger age of onset, M1 macrophage fraction in tumor microenvironment, and, in specific cancers, increased tumor mutation burden.

**Conclusions:** Our findings will help better stratification of individuals at high cancer risk, as well as the characterization of the influence of their personal germline RDV load on age of diagnosis, tumor microenvironment and mutational burden. These high-risk individuals may benefit from increased surveillance, earlier screening and prevention efforts, and treatments that exploit their tumor characteristics, improving prognosis.

## Background

Inherited genetic variants play an important role in cancer susceptibility. These variants are associated with disease risk in a spectrum, from common variants that tend to have weak effects, to rare variants (<0.5% allele frequency) with often large effects^1,2^. In fact, some studies suggest that greater than 95% of variants predicted to be functionally important are rare^3,4^. In addition to their high penetrance, rare variants are also abundant^3^. Studies in cancer prone families were the first to identify rare, deleterious variants (RDVs) with statistically significantly elevated cancer risk, and these RDVs are currently used in cancer genetics clinics to assess personal cancer risk^5^. The vast majority of genes identified this way are in DNA damage repair (DDR) genes. Consistent with this, we have recently reported that RDVs in *Fanconi Anemia* (FA) genes increase risk for lung squamous cell carcinoma^6^ and in the *ATM* gene increase risk for lung adenocarcinoma^7^.

While genome-wide association studies (GWAS) have identified multiple susceptibility loci as determinants of increased cancer risk in relatively large cohorts, these studies examine associations with common variants only. More recently, next generation sequencing studies by large consortia have produced and aggregated data from thousands of germlines and matched tumors. These studies have revealed many germline risk variants^8–12^, and provide a rich resource for investigating the association of RDVs with cancer risk^6,7,9,10^. To better characterize the roles of RDVs in cancer risk and other cancer-related outcomes, it is paramount to comprehensively assess RDVs in such cancer germline datasets. This will enable the development of predictive tools and precision preventive strategies for the clinic.

Towards this end, we have performed the largest to date jointly processed cancer case-control germline sequencing data analysis, aggregating existing whole-exome sequencing (WES) datasets on 15,709 participants, where the cases span 24 different cancers from The Cancer Genome Atlas, TCGA^13^. As the low frequencies of RDVs make genome-wide discovery difficult without large cohorts, we investigated their roles at gene and gene-set levels. While TCGA germline RDVs have been categorized previously^10^, this is the first study to report results where variants are jointly processed together with a matching control cohort. The joint processing has enabled us to study the associations between the total number of RDVs each individual has with cancer risk, or personal *RDV load*, and also with age of disease onset and disease progression. Importantly, this is also the first study to replicate the pan-TCGA RDV findings in an independent case-control cohort, where we utilized WES data on 7,771 participants from the Icahn School of Medicine at Mount Sinai (ISMMS) BioMe Biobank. Overall, our multi-scale analysis results first confirm known associations in TCGA^9^ and provide novel observations on the associations of germline RDVs in specific genes and gene-sets with cancer risk. We show in the two independent cohorts that individuals with germline RDV load in specific cancer genes are at higher risk for cancers in a dose-dependent manner. Finally, we show that germline RDV load in specific gene-sets is a potential biomarker for younger age of disease onset, tumor mutational burden (TMB), and characteristics of the tumor immune microenvironment (TME).

## Methods

### Data Sources

For discovery, we used case data from TCGA and control data from twelve population-based studies in the database of Genotypes and Phenotypes (dbGaP) (http://www.ncbi.nlm.nih.gov/gap). Briefly, we downloaded TCGA germline WES bam files from National Cancer Institute Cancer Genomics Hub (cgHub), a predecessor to the Genomic Data Commons which is no longer online. We extracted control fastq files from the NCBI Short Read Archive (SRA) for dbGaP studies listed in Supplementary Table S1. For replication, we used the exome calls from BioMe Biobank^14^ of Icahn School of Medicine at Mount Sinai (ISMMS).

### Study Cohorts

For the discovery cohort, we realigned and jointly called a case-control cohort of 8,321 TCGA cases and 7,388 dbGaP controls. Cases included participants with 24 different cancer types of different sample sizes, as listed in Supplementary Table S2. For the control samples, when the underlying dbGaP study was a case-control study focused on a disease, we only included control individuals who did not have that disease in question.

For the validation cohort, we utilized available datasets from ISMMS BioMe^14^, which is an electronic health record (EHR) linked biobank. BioMe encompasses a wide array of phenotypic and genetic data from the diverse population of ISMMS patients with a vast spectrum of medical disorders. WES data (Illumina v4 HiSeq 2500) already exist for 30,813 BioMe participants^14^. Focusing on BioMe participants with existing WES data, we identified cancer patients and healthy controls based on their International Classification of Diseases (ICD)-9 and ICD10 codes^15^, which led to 1,571 cases and 6,200 controls of European ancestry (filtering details below). The clinical characteristics of the cohorts are listed in Table 1. The study design is provided in Figure 1.

**Table 1:** Characteristics of samples in the case-control study cohorts.

| Variables |  | TCGA-dbGaP Cohort |  | BioMe Cohort |  |
| --- | --- | --- | --- | --- | --- |
|  |  | Cases<br>(6,371) | Controls<br>(6,647) | Cases<br>(1,571) | Controls<br>(6,200) |
| Gender | Male | 3099<br>(48.64%) | 4034<br>(60.69%) | 690<br>(43.92%) | 3150<br>(50.81%) |
|  | Female | 4034<br>(60.69%) | 2610<br>(39.27%) | 881<br>(56.08%) | 3050<br>(49.19%) |
|  | Missing | 42<br>(0.66%) | 3<br>(0.05%) | 0 | 0 |
| Age | Mean (yrs) | 60.20 | 57.54 | - | - |
|  | Unknown | 86<br>(1.35%) | 4509<br>(67.84%) |  |  |
| Smoking | Never | 448<br>(7.03%) | 322<br>(4.84%) | 1391<br>(88.54%) | 5418<br>(87.39%) |
|  | Yes | 1491<br>(23.40%) | 746<br>(11.22%) | 174<br>(11.08%) | 744<br>(12.00%) |
|  | Unknown | 4432<br>(69.57%) | 5579<br>(83.93%) | 6<br>(0.38%) | 38<br>(0.61%) |

**Figure 1:**
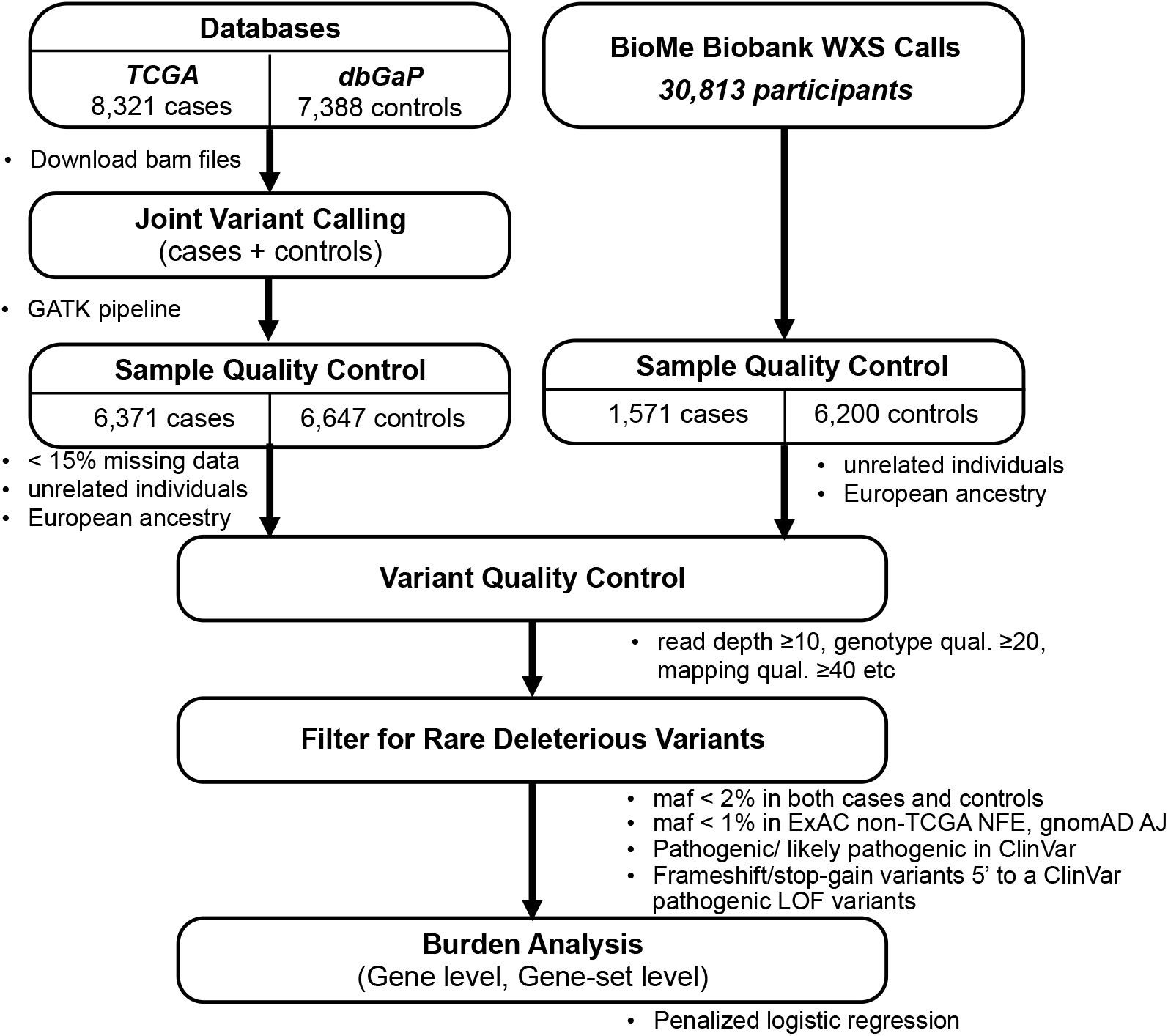
Study design on burden analysis.

### Discovery Cohort Variant Discovery

We first realigned and jointly called all case and control germline WES datasets using GVCF-based best practices for the Genome Analysis Toolkit (GATK, https://www.broadinstitute.org/gatk/) as implemented in a custom pipeline at the Icahn School of Medicine at Mount Sinai (ISMMS)^16^ and previously^6,17,18^. Briefly, we independently aligned all samples to human genome build GRCh37 with BWA^19^, performed indel realignment, duplicate marking and base quality score recalibration using GATK and Picard, and finally called to a GVCF file with HaplotypeCaller. In the joint calling step, which consisted of calling variants from GVCF files and variant quality score calibration with GATK, we only included samples for which over 75% of the exome was callable (depth >= 20, mapping quality >= 10, base quality >= 20) and for which there was no evidence of contamination (VerifyBamID < 3%).

### Discovery Cohort Sample QC

We first removed samples with 15% or more missing data. To filter samples that are duplicates or from first or second degree relative pairs, we performed relatedness analysis with KING software^20^ and then removed a sample from each such pair that had the highest fraction of missing data. To remove any bias that may arise due to systematic ancestry-based variations in allele frequency differences between cases and controls (i.e. population stratification) we used Principal Component Analysis (PCA). Briefly, we first removed indels and rare variants (defined by <5% of minor allele frequency, MAF), using 1000 Genomes dataset^21^ and The Ashkenazi Genome Consortium (TAGC, https://ashkenazigenome.org) as reference. For the remaining variants, we performed Linkage Disequilibrium (LD) pruning, filtered for a call rate of at least 0.99, and performed PCA with smartpca using EIGENSOFT 5.0.1 software. We filtered for the least ancestry-based variation by focusing our downstream analyses on the largest set of case-control individuals clustered within the PCA plot by examining the PCA plot and selecting thresholds on PC1 and PC2 to corresponded to individuals of European Ancestry (EA); analogous to flow cytometry we call this approach “gating”. The PCA plots along with the gated region are shown in Figure 2. To adjust for population-level differences, we used the first two principal components from PCA of the gated individuals as covariates in the burden analyses. After sample QC, 6,371 cases and 6,647 controls remained.

**Figure 2:**
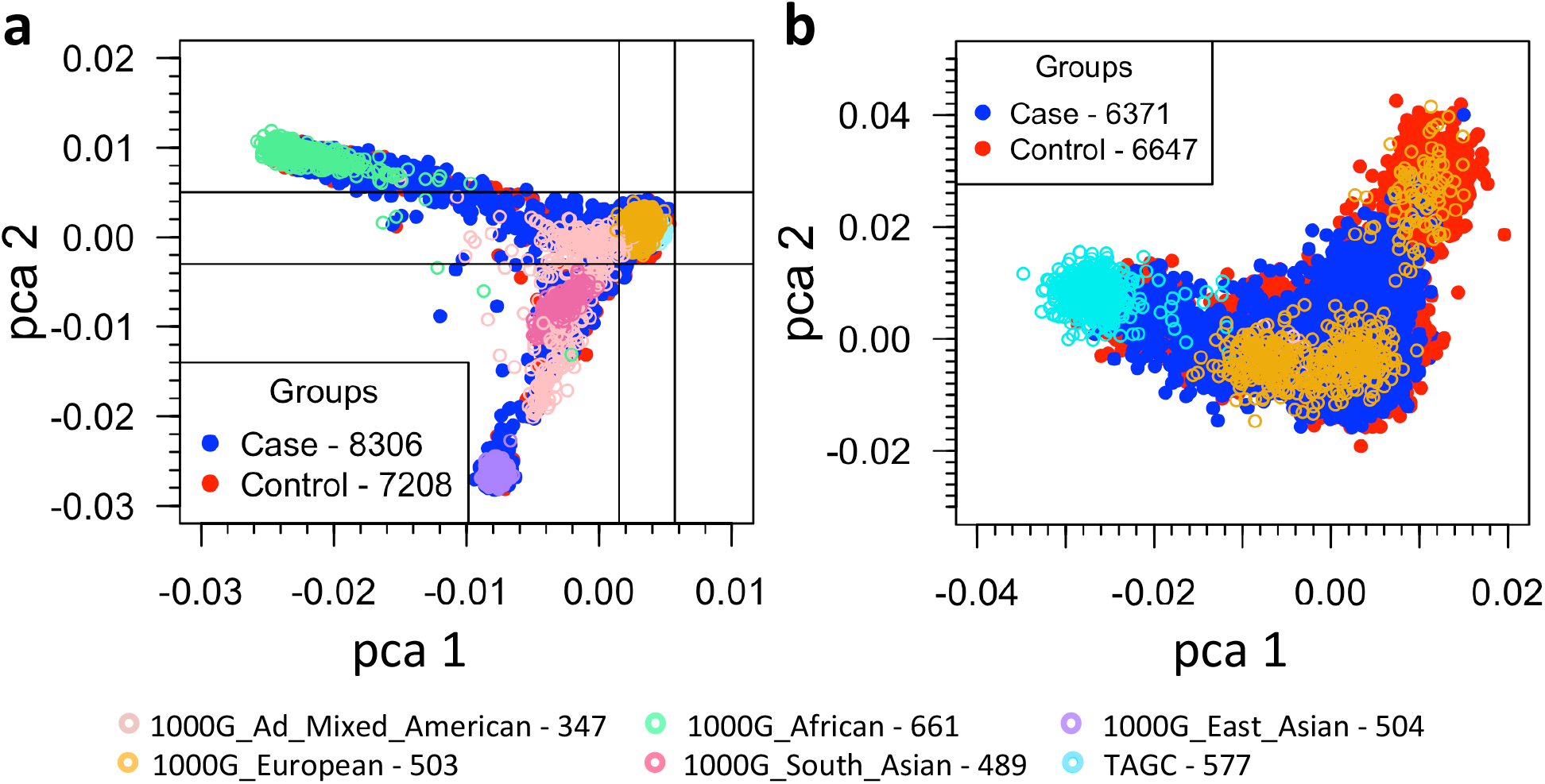
Principal Component Analyses (PCA) of the study cohort and the gated study cohort. PCA based on common SNPs (MAF > 0.05) showing the top two principal components of **a)** the study cohort together with 1000 Genomes and TAGC samples and of **b)** the gated samples from the study cohort with European ancestry (6371 cases and 6647 controls).

### Discovery Cohort Variant-level QC

For participants that passed PCA gating, we focused on ensuring high-quality genotype/variant calls for analysis. For this purpose, we filtered for variants with: read genotype quality ≥20; read depth ≥10; allelic depth of alternate allele ≥4; sites with: quality score ≥50; quality by depth score ≥2; mapping quality ≥40; read position rank sum >–3; mapping quality rank sum >–10 and variant tranche <99%. For heterozygous genotypes, we filtered for alternative allele ratio between 0.30 and 0.70. To reduce any differences between samples in cases and controls, we kept sites that have differential missing variant fraction ≤ 0.05 between the cases and controls. Finally, we kept sites with ≥88% of data (in both cases and controls).

### Validation Cohort Participant Selection

Next, to replicate the discovery cohort findings within an ancestry-matched validation cohort, we focused on those of EA in BioMe. Briefly, we used the PCA performed on the common variants^22^ and gated for EA individuals based on the first two principal components which captured the majority of the variance, resulting in 10,784 BioMe participants of (Supplementary Figure S1). Next, we also ensured we used data from unrelated participants up to second degree. Finally, we identified participants with cancer based on their ICD9 and ICD10 codes available within BioMe^14^. Specifically, to avoid any false positives, we filtered for participants who had ICD9/10 codes related to cancer at least twice in their diagnosis files on separate dates. To avoid any conflicts in categorizations, we also removed participants with the following diagnoses (or diagnoses of similar nature): benign neoplasms, neoplasm of uncertain behavior and genetic susceptibility to malignant neoplasm. The complete list of ICD9/10 codes used for classification and elimination of cancer diagnosis are listed in Supplementary Table S3. We considered all other participants who were unaccounted for in the above categories as controls. These led to 1,571 cases and 6,200 control participants of EA in the BioMe cohort.

### Validation Cohort Data Generation and Variant QC

BioMe WES data generation and QC steps have been discussed in detail previously^14^. Briefly, we filtered out sites with missingness >0.02 and biallelic sites with allele balance (<0.3 or > 0.8). Additionally, to be consistent with the discovery cohort variant QC, we filtered for variants with: read genotype quality ≥20; read depth ≥10; allelic depth of alternate allele ≥4; sites with: quality score ≥50; quality by depth score ≥2; mapping quality ≥40; read position rank sum >–3; mapping quality rank sum >–10; differential missing variant fraction ≤ 0.05 between the cases and controls and site missingness < 12% (in both cases and controls).

### Variant Filtering (both cohorts)

After sample and variant QC, we focused on rare, deleterious variants (RDVs) with known pathogenicity. To filter out common polymorphisms, we removed any variant present in both case and control cohorts at MAF > 2% or in Exome Aggregation Consortium (ExAC)^23^ non-TCGA Non-Finnish European population at MAF >1% or in Genome Aggregation Database (gnomAD)^24^ Ashkenazi Jewish population at MAF > 1%. We considered variants that pass these filters to be rare. We then filtered the remaining variants for functional impact based on those present in the ClinVar database^25^ using the Annovar tool (http://annovar.openbioinformatics.org). We considered a variant to be deleterious if: (i) it is listed as pathogenic/likely pathogenic in ClinVar; or (ii) it is a frameshift or stopgain variant located 5’ of a variant described to be a pathogenic LOF variant in ClinVar (nonsense and frameshift).

## Statistical Analysis

### Background variation correction

To test for possible background variation between cases and controls, we calculated the tally of rare autosomal synonymous variants per each study participant. We defined synonymous variants as rare at Exac MAF ≤0.005% and cohort MAF ≤0.05%. Supplementary Figure S2 provides the distribution and background variation statistics of genes with rare synonymous variants between the cases and controls in both cohorts. We accounted for differences in background variation by using the number of genes with rare synonymous variants of each individual as a covariate during the burden analyses.

### Gene level RDV burden analyses

Next, to evaluate the cumulative effects of multiple RDVs in each gene, and thereby increase the statistical power to identify cancer risk genes, we performed gene level RDV burden tests. Briefly, we performed aggregate RDV burden for each gene using Penalized Logistic Regression Analysis (PLRA), using the logistf package in R (https://cran.r-project.org/web/packages/logistf/index.html). To adjust for background variation, we used the number of genes with rare synonymous variants as a covariate for each individual in both cohorts. Additionally, in the discovery cohort we used the first two principal components as covariates to adjust for population difference.

### Gene-set level RDV burden analyses

Next, we evaluated the RDV burden of gene-sets that typically play key roles in cancer risk and progression: i) known cancer predisposition genes^26^ ; ii) DNA damage repair genes^27,28^ ; iii) somatic cancer driver genes^29^ and iv) Fanconi Anemia genes^30^. We provide a complete list of genes in these gene-sets in Supplementary Table S4. For burden analyses, we used PLRA and considered all participants with at least one RDV in a gene within the considered gene-set. Furthermore, for an unbiased data-driven exome-wide gene-set analysis, we also tested all gene-sets (17,810) in Molecular Signatures Database (MSigDB) (software.broadinstitute.org) using the same RDV burden approach.

### Gender effects

The gender breakdown of participants for each cancer type in TCGA is provided in Supplementary Table S5. To study the effect of gender on burden analyses, we first removed samples with missing gender data (Supplementary Table S5) and then used PLRA with gender as an additional covariate. Resulting ORs and *p* values of all gene-sets are in supplementary Figure S3. Please note that we did include gender-biased cancer subtypes breast, prostate, ovarian, cervical and uterine (endometrial and sarcoma) in the pan-cancer analysis of gender effects.

### Germline RDV load effects

Next, we asked whether the accumulation of personal germline RDVs, or ‘RDV load” of an individual impacts their personal cancer risk. Towards this end, we first divided the discovery cohort participants based on their RDV loads into three groups: participants with i) no RDVs; ii) one RDV; and iii) more than one RDV. We then tested and compared the association of germline RDV load with age of diagnosis between these groups using the Mann-Whitney U test. Next, we asked whether the germline RDV load impacts tumor immune microenvironment (TME). For this purpose, we used existing datasets^31^ on the relative fraction of 22 different immune cell types within TME across TCGA cancers, as estimated by the CIBERSORT tool^32^, which included 5,917 cases. To obtain the total cell fraction in tissue, we multiplied the relative immune cell fractions with leukocyte fraction^31^. We then compared the immune cell fractions between the groups using the Kruskal-Wallis test. Next, to study the effect of M1 macrophages on survival, we used Cox proportional hazards regression model. Finally, to study the effect of germline RDVs on tumor mutation burden (TMB) between the groups, we used Mann-Whitney U test. To calculate the TMB, we used the publicly available TCGA somatic mutations MAF file (mc3.v0.2.8.PUBLIC.maf.gz) which included 6,225 cases. We used the TMB definition as the total number of somatic, missense, nonsense, frameshift/inframe mutations per megabase (Mb) of genome examined, with 38 Mb as an estimate of exome size.

## Results

### Joint case-control variant analysis identified sites of rare, deleterious variants (RDVs) and their genotypes across the cohort

To enable case-control analyses (Figure 1) while avoiding biases potentially introduced by different calling algorithms, we realigned and jointly called variants in the germline WES data from 8,321 TCGA cases (Supplementary Table S2), together with 7,388 controls from dbGaP (Supplementary Table S1). To reduce confounding due to population stratification, we focused on those participants in the discovery cohort who comprised the largest group by ancestry, which genetically appeared to be of European ancestry (Figure 2). After sample and variant QC (see methods), we observed 941,609 variants (Supplementary Table S6) in 17,507 genes across the autosomes and X chromosomes of 6,371 cases and 6,647 controls (clinical characteristics in Table 1). We note that recently an independent pan-cancer analysis of TCGA cases^10^ (‘case-only analysis’), which called each case sample separately (rather than jointly calling variants across all samples) and used the union of several calling software packages for variant identification. Unlike that case-only approach, our joint calling approach can distinguish between instances where a variant site is wild type and where it does not have enough sequence coverage to make a call. Overall, we identified 7,241 RDVs (MAF< 2% in cases or controls) across 1,787 genes in the discovery cohort. Similarly, focusing on 1,571 cases and 6,200 control participants of European ancestry in the validation cohort, we identified 5,766 RDVs in 1,814 genes.

### Gene-set level burden analysis revealed that RDVs in cancer predisposition, DNA damage repair, *Fanconi Anemia* and somatic cancer driver gene-sets are associated with cancer risk

Though we performed tests at the levels of individual genes, we did not find any significant, replicable associations beyond the well-known association of *BRCA1/2* with breast and ovarian cancer risk (Supplementary Table S7). Instead, we hypothesized that collapsing RDVs at the gene-set level would be more powerful and enable better understanding of risk-associated biological processes. Therefore, we compared the RDV burden (Table 2) in *a priori* defined gene groups (Supplementary S4). We first tested the set of 94 genes in the TruSight Cancer Gene panel^26^, which is often used in genetic testing clinics, and observed statistically significant RDV burden across cancers in the discovery cohort (OR=1.51; *p*-value= 4.58e-08, 95% CI: 1.30-1.75), which replicated in the validation cohort (OR=1.24; *p*-value= 0.04, 95% CI: 1.01-1.51). Similarly, when we focused on 95 DNA repair genes involved in known functional DDR pathways^27^ and are known to associate with autosomal dominant CPD syndromes^28^ (Supplementary Table S4), we observed a statistically significant enrichment of RDVs in cases compared to controls in both discovery (OR=1.50; *p*-value= 8.30e-07, 95% CI: 1.28-1.77) and validation cohorts (OR=1.59; *p*-value= 1.17e-04, 95% CI: 1.26-2.00). Next, based on the Knudson’s second-hit hypothesis^33^ that a somatic mutation could be the second hit of a germline RDV (i.e. a biallelic mutation), we tested the germline RDV burden on 299 known somatic cancer driver (SCD) genes^29^. We again observed a statistically significant burden of RDVs across cancers in cases vs. controls in the discovery cohort (OR=1.46; *p*-value= 4.04e-06; 95% CI, 1.24-1.72), which replicated in the validation cohort (OR=1.72; *p*-value= 2.00e-06; 95% CI, 1.38-2.14).

**Table 2.** Geneset-level rare, deleterious variant (RDV) burden in the study cohorts.

|  | TCGA-dbGaP Cohort |  | BioMe Cohort |  |
| --- | --- | --- | --- | --- |
|  | Cases<br>(6,371) | Controls<br>(6,647) | Cases<br>(1,571) | Controls<br>(6,200) |
|  | 94 Cancer predisposition genes |  |  |  |
| # Variants | 274 | 186 | 88 | 192 |
| # Genes | 57 | 48 | 35 | 51 |
| # Unique individuals | 464 (7.28%) | 326 (4.90%) | 136 (8.66%) | 439 (7.08%) |
| OR (*p*-value) (95% CI) | 1.51 (4.58e-08) [1.30-1.75] |  | 1.24 (0.04) [1.01-1.51] |  |
|  | 95 DNA damage repair genes |  |  |  |
| # Variants | 254 | 181 | 82 | 160 |
| # Genes | 41 | 36 | 29 | 35 |
| # Unique individuals | 374 (5.87%) | 269 (4.05%) | 108 (6.87%) | 273 (4.40%) |
| OR (*p*-value) (95% CI) | 1.50 (8.30e-07) [1.28-1.77] |  | 1.59 (1.17e-04) [1.26-2.00] |  |
|  | 299 Somatic cancer driver genes |  |  |  |
| # Variants | 231 | 154 | 88 | 169 |
| # Genes | 59 | 52 | 40 | 59 |
| # Unique individuals | 377 (5.92%) | 272 (4.09%) | 125 (7.96%) | 294 (4.74%) |
| OR (*p*-value) (95% CI) | 1.46 (4.04e-06) [1.24-1.72] |  | 1.72 (2.00e-06) [1.38-2.14] |  |
|  | 22 Fanconi Anemia genes |  |  |  |
| # Variants | 115 | 58 | 32 | 69 |
| # Genes | 15 | 13 | 8 | 13 |
| # Unique individuals | 163 (2.56%) | 85 (1.28%) | 52 (3.31%) | 103 (1.66%) |
| OR (*p*-value) (95% CI) | 2.05 (6.14e-08) [1.58-2.70] |  | 2.01 (1.07e-04) [1.42 - 2.80] |  |

Based on our earlier findings^6^ that RDVs in *Fanconi Anemia (FA)* genes increased risk for squamous lung cancer (LUSC), we also investigated the burden of FA (subset of 22 DDR genes) RDVs in other tissues and pan-cancer. Consistent with our LUSC findings, we observed an increased RDV burden cross-cancers in cases vs. controls in both discovery (OR=2.05; *p*-value= 6.14e-08; 95% CI: 1.58-2.70) and validation (OR=2.01; *p*-value= 1.07e-04; 95% CI: 1.42-2.80) cohorts. In the discovery cohort, this signal was significantly driven by cancers of the breast, bladder, stomach and ovary (Supplementary Table S8). Furthermore, the 9 *FA* core complex genes and 11 *FA* genes involved in DNA repair both had a statistically significant signal cross-cancers in the discovery cohort (OR=1.93; *p*-value= 0.02; 95% CI: 1.10-3.49 and OR=2.12; *p*-value= 2.15e-06; 95% CI: 1.54-2.93, respectively). The finding on 11 *FA* genes involved in DNA repair further replicated in the validation cohort (OR=2.48; *p*-value= 3.14e-06; 95% CI: 1.71-3.56), while on 9 *FA* core complex genes trended in the expected direction (OR=1.18; *p*-value= 0.74; 95% CI: 0.41-2.88). Next, to make sure the signals we observed in these gene-sets (CPD, DDR, SCD and *FA*) were not solely driven by *BRCA1/2*, as a sensitivity analysis we removed *BRCA1/2* and repeated the burden analyses, and observed that these gene-sets were still significant (Supplemental Table S9). Note that while we further tested for the confounding effects of gender for all gene-sets pan-cancer, we did not observe a noticeable difference in the OR results for any gene-set (Supplementary Figure S3). We provide the gender distribution in Supplementary Table S5.

### Data-driven analysis of 17,810 gene-sets in MSigDB again identified RDV burden in *Fanconi Anemia* genes associated with increased cancer risk

Next, to ensure we did not miss any additional gene-sets with significant RDV burden in cases vs. controls, we performed a data-driven exome-wide gene-set analysis. For this purpose, we tested all gene-sets (17,810) in Molecular Signatures Database (MSigDB) (software.broadinstitute.org; Supplementary Table S10). Consistent with our observations on pre-selected gene-sets, the strongest signal was in *Fanconi Anemia* pathway genes from REACTOME (*p*=1.4e-11; OR=2.54; 95%CI = 1.92-3.40) in the discovery cohort, which replicated in the validation cohort (*p*=1.2e-05; OR=2.21; 95%CI = 1.56-3.09). Significant signals after correcting for multiple testing were also found in other cancer-related pathways, most of which include the strongest DDR genes. Of note in the discovery cohort, we observed significant risk association for RDV burden in the set of genes targeted by the eukaryotic translation initiation factors *EIF4EBP1* and *EIF4EBP2* (OR=1.43; *p*-value=4.4e-05; 95% CI = 1.20-1.69). While we did not observe significant association in the validation cohort, we still observed higher frequency with the same direction of effect in cases with RDVs compared to controls (OR=1.14; *p*-value=0.39; 95% CI = 0.85-1.51).

### Germline *RDV load* in key cancer genes is a potential marker for increased cancer risk, younger age of disease onset, altered tumor immune microenvironment and increased tumor mutation burden

RDVs in cancer are usually considered in a binary context; a patient either has an RDV in a gene of interest or does not. Given that even when the penetrance of RDVs in cancer risk is high, it is not absolute (i.e. some individuals with RDVs in known cancer predisposition genes will never develop cancer), we hypothesized that the accumulation of RDVs within a set of related genes in an individual could increase their cancer risk. We term this concept “germline RDV load”, and hypothesized that increased germline RDV load in particular gene-sets will elevate an individual’s risk of cancer. The premise of this hypothesis is that the RDVs we are considering damage, but do not fully break, a particular pathway; therefore, additional RDVs in the same pathway add to the damage and increase cancer risk further. To test this hypothesis, we evaluated the associations between the participant-level RDV load within the CPD, DDR, SCD and FA gene-sets (Supplementary Table S4) and cancer risk (Table 3). For each gene-set, we divided the participants into three participant groups: i) participants with no RDVs; ii) participants with RDVs in only one gene; and iii) participants with RDVs in two or more genes. Consistent with our hypothesis on RDV load, for each studied gene-set, we observed a greater association with cancer risk for participants that had higher number of genes with germline RDVs, as shown in Table 3 and Figure 3 (except for FA due to the limited number of genes in this gene-set). Notably, all our RDV load findings replicated in the validation cohort.

**Table 3.** Geneset-level rare, deleterious variant (RDV) burden in the study cohorts based on the number of genes with RDVs.

|  | TCGA-dbGaP Cohort |  | BioMe Cohort |  |
| --- | --- | --- | --- | --- |
|  | Cases<br>(6,371) | Controls<br>(6,647) | Cases<br>(1,571) | Controls<br>(6,200) |
|  | 94 Cancer predisposition genes |  |  |  |
| # Indiv. without RDVs | 5907 | 6321 | 1435 | 5761 |
| # Indiv. with RDVs in 1 gene | 452 | 323 | 128 | 426 |
| # Indiv. with RDVs in ≥ 2 genes | 12 | 3 | 8 | 13 |
| Comparing Indv with RDV in 1vs0<br>OR (*p*-value) (95% CI) | 1.48 (1.68e-07) [1.28-1.72] |  | 1.20 (0.08) [0.98-1.47] |  |
| Comparing Indv with RDV in ≥2vs0<br>OR (*p*-value) (95% CI) | 3.31 (0.03) [1.10-13.02] |  | 2.55 (0.04) [1.03-5.94] |  |
|  | 95 DNA damage repair genes |  |  |  |
| # Indiv. without RDVs | 5997 | 6378 | 1463 | 5927 |
| # Indiv. with RDVs in 1 gene | 367 | 269 | 103 | 268 |
| # Indiv. with RDVs in ≥ 2 genes | 7 | 0 | 5 | 5 |
| Comparing Indiv with RDV in 1vs0<br>OR (*p*-value) (95% CI) | 1.47 (2.80e-06) [1.25-1.74] |  | 1.55 (3.75e-04) [1.22-1.95] |  |
| Comparing Indiv with RDV in ≥2vs0<br>OR (*p*-value) (95% CI) | 15.13 (0.007) [1.82-1968.26] |  | 3.96 (0.03) [1.17-13.41] |  |
|  | 299 Somatic cancer driver Genes |  |  |  |
| # Indiv. without RDVs | 5994 | 6375 | 1446 | 5906 |
| # Indiv. with RDVs in 1 gene | 366 | 267 | 123 | 288 |
| # Indiv. with RDVs in ≥ 2 genes | 11 | 5 | 2 | 6 |
| Comparing Indiv with RDV in 1vs0<br>OR (*p*-value) (95% CI) | 1.44 (9.56e-06) [1.23-1.70] |  | 1.73 (2.01e-06) [1.39-2.15] |  |
| Comparing Indiv with RDV in ≥2vs0<br>OR (*p*-value) (95% CI) | 2.19 (0.12) [0.82-6.63] |  | 1.56 (0.56) [0.29-6.12] |  |
|  | 22 Fanconi Anemia Genes |  |  |  |
| # Indiv. without RDVs | 6208 | 6562 | 1519 | 6097 |
| # Indiv. with RDVs in 1 gene | 163 | 85 | 51 | 101 |
| # Indiv. with RDVs in ≥ 2 genes | 0 | 0 | 1 | 2 |
| Comparing Indiv with RDV in 1vs0<br>OR (*p*-value) (95% CI) | 2.05 (6.14e-08) [1.58-2.70] |  | 2.01 (1.24e-04) [1.42-2.80] |  |
| Comparing Indiv with RDV in ≥2vs0<br>OR (*p*-value) (95% CI) | - |  | 2.38 (0.42) [0.22-17.93] |  |

**Figure 3:**
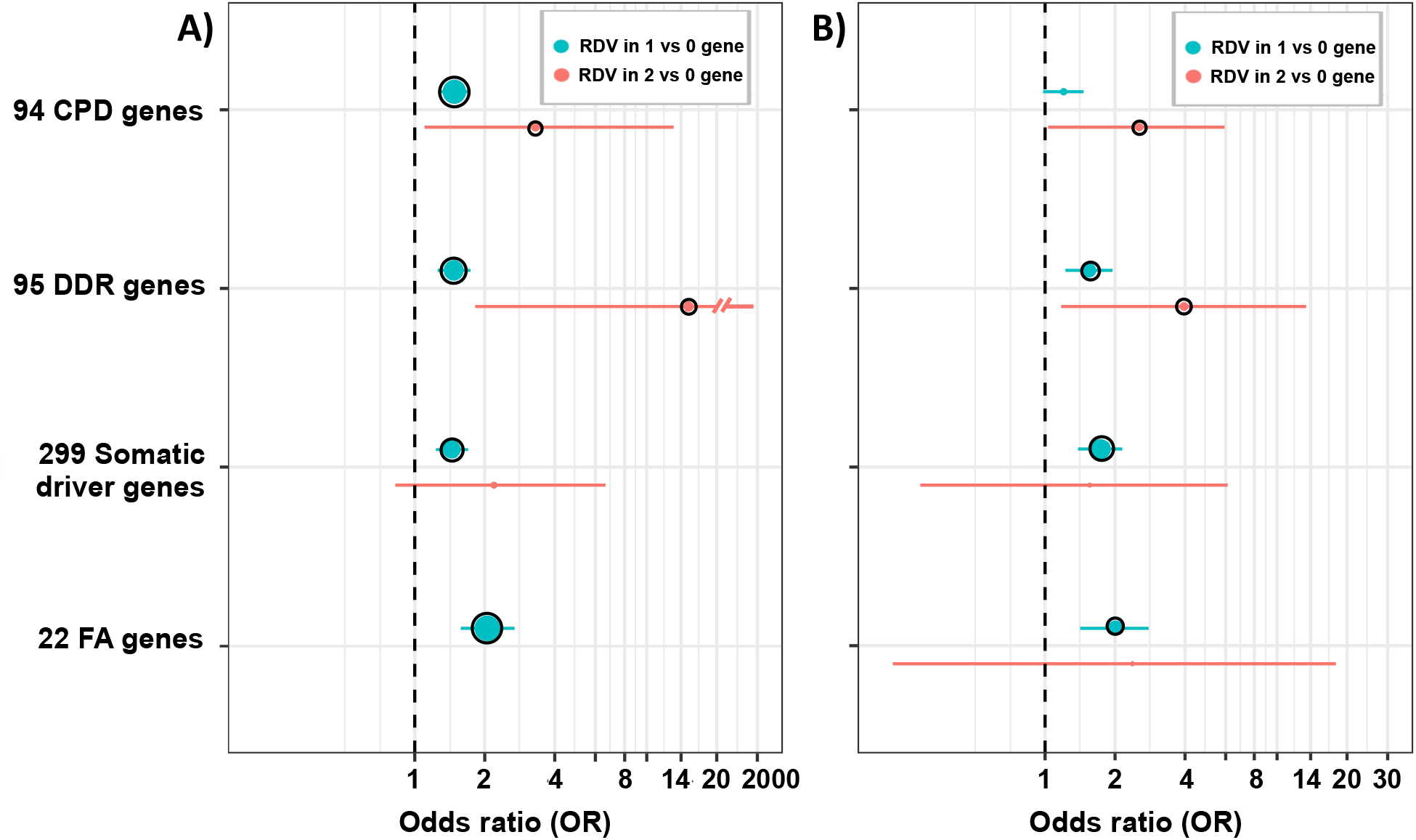
Cancer risk based on RDV load. **A)** Discovery cohort; **B)** Validation cohort. The whiskers span the 95% confidence interval for OR values. The black circle outline indicates significant burden *p* ≤ 0.05

We further hypothesized that a higher RDV load in these gene-sets will result in an earlier age of diagnosis. To test this hypothesis, we used the detailed clinical information available on the discovery cohort of TCGA participants. We evaluated the associations between each participant’s germline RDV load with their age at cancer diagnosis, and observed that those participants with higher germline RDV load indeed exhibited a statistically significant early age of diagnosis (Figure 4).

**Figure 4.**
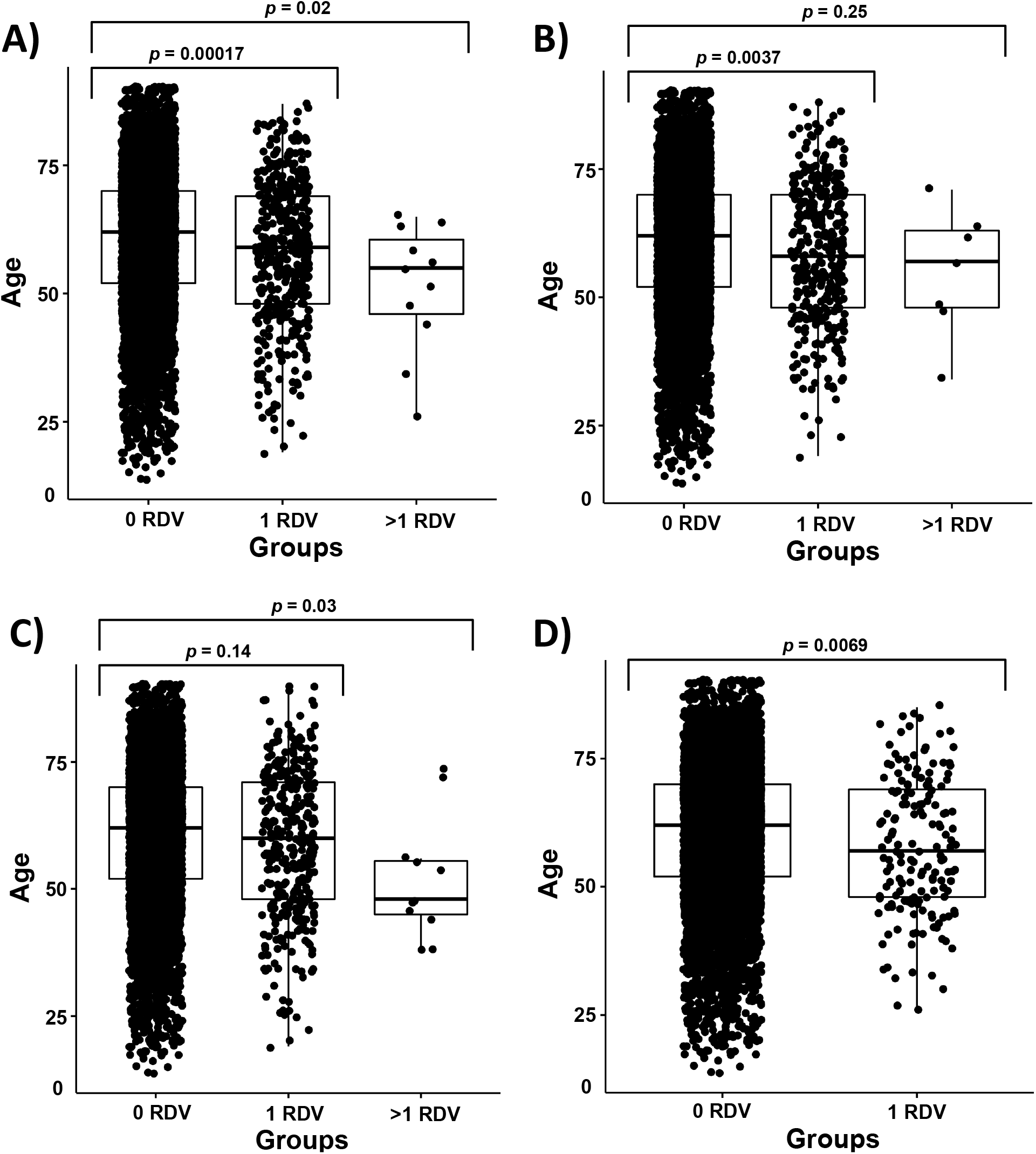
Comparison of age of diagnosis based on germline RDV load in TCGA cases. **A)** Cancer predisposition genes **B)** DNA damage repair genes **C)** Somatic cancer driver genes **D)** Fanconi Anemia genes

Next, in light of recent findings on the impact of germline variants on tumor immune microenvironment (TME) in various cancers^34–36^, we tested the association of increased germline RDV load in CPD, DDR, SCD and *FA* gene-sets with TME immune cell fractions. Briefly, to avoid any potential biases from B-cell immune signatures in blood cancers, we focused on 6,277 participants with solid tumors in TCGA by excluding 94 TCGA participants with hematological malignancies. We then utilized previously reported annotations on the 22 infiltrating immune cell types on solid tumors of TCGA participants^31^, based on analysis by the CIBERSORT tool that used their tumor RNA-sequencing data^32^. Next, for each participant, we tested the association of tumor immune cell fraction with their germline RDV load in the CPD, DDR, SCD and *FA* gene-sets (Supplementary Figure S4, Supplementary Table S11). Remarkably, we observed that participants with a germline RDV in CPD, DDR or *FA* gene-sets (but not SCD gene-set) developed tumors with a statistically significantly higher fraction of M1 Macrophages (Figure 5), compared to participants with no RDV. This signal was mostly driven by the increased levels of the chemokine ligands *CLXCL10/11*. We provide the complete set of results in Supplementary Table S12. We next asked whether increased M1 macrophages in TME of participants with solid tumors associated with survival (Supplementary Table S13). We observed that those participants with increased M1 macrophages exhibited worse survival in brain lower grade glioma, kidney renal clear cell carcinoma and kidney renal papillary cell carcinoma.

**Figure 5.**
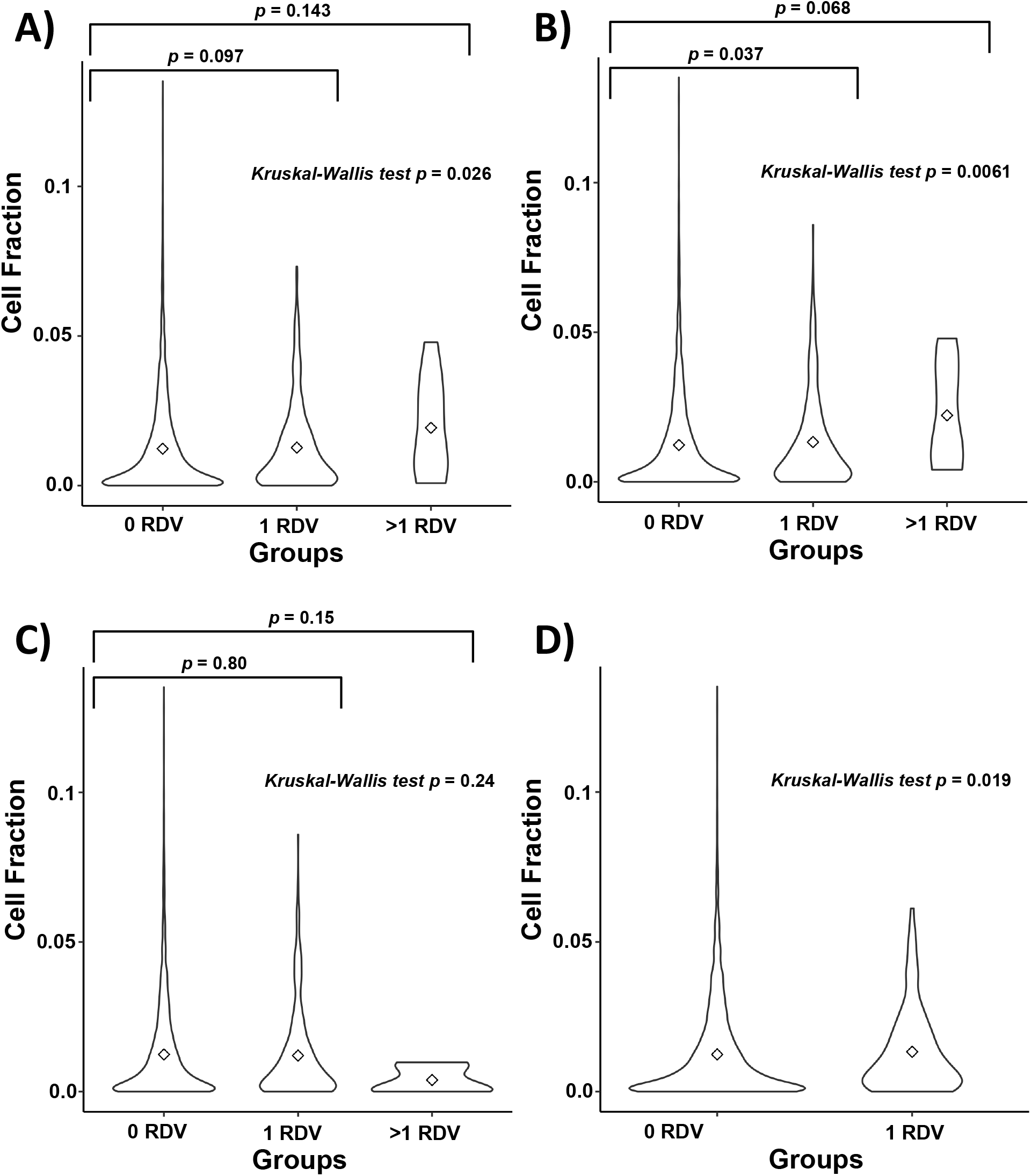
Comparison of M1 Macrophages cell fraction in tumor based on germline RDV load in TCGA cases. **A)** Cancer predisposition genes **B)** DNA damage repair genes **C)** Somatic cancer driver genes **D)** Fanconi Anemia genes

Finally, since tumor mutation burden (TMB) is currently being investigated as a biomarker for durable response to life-extending cancer immunotherapies^37^, we also tested the association of germline RDV load in CPD, DDR, SCD and *FA* gene-sets with TMB for each cancer type with solid tumors (Supplementary Table S14). We observed the strongest TMB association in breast cancers, with germline RDV load in *FA* genes (p=0.00052). Overall, germline RDVs associated with TMB in: i) DDR, CPD and SCD genes in colon cancer; ii) DDR genes in kidney clear cell and cervical cancers; iii) CPD genes in kidney papillary cell cancers; iv) SCD genes in stomach and ovarian cancers; and v) *FA* genes in breast, cervical, ovarian and stomach cancers. This signal for *FA* genes was mostly driven by *BRCA1/2*; when we removed *BRCA1/2* and reperformed the analysis, we still observed associations of germline RDV load in *FA* genes and TMB for stomach cancer. Details are in Supplementary Table S14.

## Discussion

Here, we have performed the largest-to-date case-control WES study to identify germline rare, deleterious variants (RDVs) that associate with pan-cancer risk and by far the first pan-cancer WES study that replicates results in an independent case-control cohort. Uniquely, in addition to studying the presence of RDVs in particular genes or gene-sets as a dichotomous variable, analyzing the cases together with thousands of controls enabled us to investigate whether an increased load of germline RDVs in individuals increased their personal cancer risk and tumor characteristics.

We first performed gene-level analyses as positive controls of our approach. As the low frequencies of RDVs make genome-wide discovery difficult without large cohorts, aggregating RDVs at gene and biologically related functional gene group levels for joint analyses increases statistical powe^38–41^ for their cumulative effect on cancer risk^6^ by reducing the multiple hypothesis burden. After collapsing all germline RDVs at gene level^42^, 501 genes associated with risk (Supplementary Table S7). Interestingly, the majority of the identified genes (457 out of 501) was not known cancer predisposition (CPD) or DNA damage repair (DDR) genes; nor were they somatic cancer driver (SCD) genes (Supplementary Table S4). Of these, the known cancer risk genes^43^ *BRCA1/2* replicated in the validation cohort. BRCA1/2 were previously established to have the highest frequency RDVs in cancer risk, and identified in earlier TCGA studies^10^, validating our approach.

Next, we took a gene-set based variant collapsing approach to examine the cumulative effects of RDVs in functionally related gene groups. Specifically, mindful of the fact that many risk variants have been shown to exhibit tissue specificity^44^, we focused on RDVs in CPD and DDR genes, which we predicted would pleiotropically associate with risk across cancer sites, due to the accumulation of mutations that fail to be properly repaired^45^. In addition, based on Knudsons’ two-hit hypothesis^33^ that a somatic mutation could be a second hit after a germline RDV, we also collapsed germline RDVs in SCD genes^29^. Finally, based on our earlier findings^6^ that RDVs in *FA* genes are associated with lung squamous risk, we also focused on RDVs in the *FA* gene-set. Remarkably, we observed increased cancer risk associated with RDVs in all four gene-sets in both discovery and validation cohorts.

Our results demonstrate the value of integrative analysis of case and control participants in germline risk variant discovery. Notably, unlike prior works^10^ that have simply compared germline variant frequencies in TCGA cancer patients to those in databases such as ExAC^23^ non-TCGA or gnomAD^24^ as controls, we have jointly called the variants in case and control participants. Doing so enabled us to examine the role of *RDV load* in cancer risk. While multiple studies have focused on the somatic mutational load of tumors, ours is arguably the first to highlight the critical importance of germline RDV load in key cancer gene-sets. The association of increased RDV load in CPD, DDR, *FA* and SCD gene-sets with increased personalized cancer risk in both discovery and validation cohorts have important implications for our understanding of how germline genetic factors govern cancer risk, and can impact the clinical management of cancer patients with one or more germline RDVs, and their families.

In addition to our results on cancer risk, we observed novel findings on the association of RDV load with age of disease onset, tumor immune microenvironment (TME) and tumor mutation burden (TMB) using TCGA matching tumor data. That individuals who have multiple RDVs in CPD, DDR or *FA* genes have statistically significantly earlier age of diagnosis than those without suggests whether starting screening and surveillance efforts at younger ages than the currently recommended guidelines for the general public is warranted for these individuals.

TME and TMB are two tumor characteristics important in immunotherapy response and prognosis. Recent studies have shown that germline variants can shape certain immune features within the TME of solid tumors^34–36^. Consistent with and complementary to these observations, our results show that the personal germline RDV load of a patient can impact their TME (see Supplementary Figure S4, Supplementary Table S11). Germline RDV load in CPD, DDR or *FA* genes exhibited the strongest association with increased levels of M1 macrophages in the TME (Figure 5). Typically, unlike T-cells which may or may not be in the TME, macrophages are generally present in tumors often as the most dominant cells, including tissue resident cells and infiltrating cells, with relative proportions varying in different patients.. The potential impact of increased macrophages on the tumor is currently somewhat controversial, and historical dichotomy between good (M1) vs. bad (M2) subsets has now been challenged by more granular data of macrophage subsets and lineage based on single-cell transcriptomics. M1 macrophages have historically been shown to be beneficial by attending tumor repair, but they can also dampen T-cell recruitment and function and regulate other aspects of tumor immunity. However, while increased levels of M2 macrophages generally associate with poor survival in tumors, the potential impact of increased M1 macrophages have so far not been associated necessarily with poor outcome. Given that TMI is known to impact responses to standard-of-care therapies, including chemotherapy, radiation and angiogenic inhibitors, and that clinical trials that examine the combinations of these treatments are currently on-going, our results warrant further investigations into better defining specific macrophage markers that may correlate with immunotherapy treatment efficacy and prognosis of individuals with high germline RDV load in CPD, DDR or *FA* genes in solid tumors.

We also observed statistically significant association of RDV load with TMB in different gene-sets for different cancer types (see Supplementary Table S14). Given that TMB has been suggested as a biomarker for durable response to cancer immunotherapies^37,46^, these results further support our findings on the potential critical importance of RDV load in these gene-sets in shaping certain tumor immune characteristics (Supplementary Table S11). For example, studying the RDV load associations per cancer type, we observed that in colon cancer, RDVs in most gene-sets we tested (CPD, DDR and SCD) were associated with increased TMB. These findings on tumor characteristics naturally lead to the question of whether the RDV load also impacts survival. Unfortunately, we had limited statistical power to answer this question directly. However, while observational, increased M1 macrophages (which correlated with RDV load) did associate with worse survival (Supplementary Table S13) in specific cancer types (brain lower grade glioma, kidney renal clear cell carcinoma and kidney renal papillary cell carcinoma). The functionality of macrophages in these histologies, in relation to antigen presentation capacity or inflammatory potential, compared to macrophages in other common tumor types, should be further explored. Further studies in larger, independent cohorts are needed to better understand the nature of interactions between germline RDV load and tumor characteristics and survival.

This study should also be considered in the context of its limitations. First, the pan-cancer analysis needs to consider the overrepresentation of rarer cancers in the TCGA data. Thus, strong association signals specific to these less common cancers in the TCGA data may be weaker when studied in a cohort reflective of the population incidences of cancers of various sites. Second, while focusing on variants annotated as pathogenic in ClinVar^25^ was necessary to ensure the clinical reliability of our results, it also restricts our analysis to those genes previously known to have a clinical impact. There are variants, such as rs11571833 (p.Lys3326Ter) in *BRCA2*, for which research studies strongly support a role in cancer risk^6,47^ but not annotated as pathogenic in ClinVar. Alternative approaches to identifying pathogenic variants will be needed to address these issues. Third, while we had well-annotated gender data available to study its potential confounding effects, such information on age and smoking was not available for all controls in the dbGaP studies and thus we were unable to investigate their potential confounding effects. Fourth, our results do not explore the interplay between specific RDVs, RDV load, and environmental and clinical exposures. Future efforts that link genetic information with epidemiological exposure and clinical information from Electronic Health Records (EHR) (e.g. blood measurements; smoking and alcohol history; viral infections, etc) will be needed to understand such interactions. This work also did not consider the role of common genetic polymorphisms in cancer risk, such as that captured by polygenic risk scores^48^. Future efforts towards understanding if and how the penetrance of RDV load will be impacted with polygenic background could be quite informative^49,50^. Finally, the statistical power of our validation was somewhat limited. As data from larger population-or hospital-based BioBanks (e.g. UKBiobank) become available, larger studies to interrogate the role of RDV load in pan-cancer and tissue-specific cancer risk will become possible.

## Conclusions

In summary, we have performed the largest-to-date case-control WES analyses to identify germline RDVs that associate with pan-cancer risk at gene and gene-set levels and validated the results in an independent cohort. Additionally, we discovered that a person’s germline RDVs load in specific gene-sets can impact their cancer risk, age of diagnosis and tumor characteristics. These gene-sets include (i) cancer predisposition genes; ii) DNA damage repair genes; iii) somatic cancer driver genes; and iv) Fanconi Anemia genes. Our findings on increased personal germline RDV load in these gene-sets will aid in identifying high-risk individuals, as well as age of disease onset and tumor characteristics. Cancer prevention and early detection in these high-risk individuals may be achieved through increased surveillance, earlier screening and other personalized targeted prevention efforts, improving prognosis.

## Data Availability

For our discovery cohort, we used case data from TCGA and control data from twelve population-based studies in the database of Genotypes and Phenotypes (dbGaP) (http://www.ncbi.nlm.nih.gov/gap). We downloaded TCGA germline WES bam files from National Cancer Institute Cancer Genomics Hub (cgHub), a predecessor to the Genomic Data Commons (https://portal.gdc.cancer.gov) which is no longer online. We extracted control fastq files from the NCBI Short Read Archive (SRA) for the following dbGaP studies:phs000209, phs000276, phs000296, phs000298, phs000424, phs000654, phs000687, phs000806, phs000876, phs000971, phs001000 and phs001101. For replication, we used the exome calls from BioMe Biobank of Icahn School of Medicine at Mount Sinai (ISMMS).

http://biomebiobank.mssm.edu:8080/biobank/service/biobankHome.jsp

## Acknowledgements

This work was supported by grants to Z.H.G from LUNGevity Foundation, Uniting Against Lung Cancer Foundation; to R.J.K. from the National Cancer Institute (R01 CA167824); and in part through the computational resources and staff expertise provided by Scientific Computing at the Icahn School of Medicine at Mount Sinai. S.G. was supported by grants U24 CA224319 and U01 DK124165. The results here are in part based upon data generated by the TCGA Research Network (https://www.cancer.gov/tcga) and datasets from dbGaP (http://www.ncbi.nlm.nih.gov/gap) through dbGaP accession numbers phs000209, phs000276, phs000296, phs000298, phs000424, phs000654, phs000687, phs000806, phs000876, phs000971, phs001000, phs001101. The authors would like to thank the participants, investigators and staff of the above studies. Additionally, the authors would also like to include the following acknowledgement statements for the dbGap studies:

***phs000209:*** MESA and the MESA SHARe project are conducted and supported by the National Heart, Lung, and Blood Institute (NHLBI) in collaboration with MESA investigators. Support for MESA is provided by contracts N01-HC-95159, N01-HC-95160, N01-HC-95161, N01-HC-95162, N01-HC-95163, N01-HC-95164, N01-HC-95165, N01-HC-95166, N01-HC-95167, N01-HC-95168, N01-HC-95169, UL1-RR-025005, and UL1-TR-000040.

This study is part of the NHLBI Grand Opportunity Exome Sequencing Project (GO-ESP). Funding for GO-ESP was provided by NHLBI grants RC2 HL103010 (HeartGO), RC2 HL102923 (LungGO) and RC2 HL102924 (WHISP). The exome sequencing was performed through NHLBI grants RC2 HL102925 (BroadGO) and RC2 HL102926 (SeattleGO). HeartGO gratefully acknowledges the following groups and individuals who provided biological samples or data for this study. DNA samples and phenotypic data were obtained from the following studies supported by the NHLBI: the Atherosclerosis Risk in Communities (ARIC) study, the Coronary Artery Risk Development in Young Adults (CARDIA) study, Cardiovascular Health Study (CHS), the Framingham Heart Study (FHS), the Jackson Heart Study (JHS) and the Multi-Ethnic Study of Atherosclerosis (MESA).

***phs000276:*** The NFBC1966 Study is conducted and supported by the National Heart, Lung, and Blood Institute (NHLBI) in collaboration with the Broad Institute, UCLA, University of Oulu, and the National Institute for Health and Welfare in Finland. This manuscript was not prepared in collaboration with investigators of the NFBC1966 Study and does not necessarily reflect the opinions or views of the NFBC1966 Study Investigators, Broad Institute, UCLA, University of Oulu, National Institute for Health and Welfare in Finland and the NHLBI.

***phs000296:*** This research used data generated by the COPDGene study, which was supported by NIH grants U01 HL089856 and U01 HL089897. The COPDGene project is also supported by the COPD Foundation through contributions made by an Industry Advisory Board comprised of Pfizer, AstraZeneca, Boehringer Ingelheim, Novartis, and Sunovion. This study is part of the NHLBI Grand Opportunity Exome Sequencing Project (GO-ESP). Funding for GO-ESP was provided by NHLBI grants RC2 HL103010 (HeartGO), RC2 HL102923 (LungGO) and RC2 HL102924 (WHISP). The exome sequencing was performed through NHLBI grants RC2 HL102925 (BroadGO) and RC2 HL102926 (SeattleGO).

***phs000298:*** The data set(s) were deposited by the ARRA Autism Sequencing Collaborative, an ARRA funded research initiative. Support for the Autism Sequencing Collaborative was provided by grants: R01-MH089208 awarded to Dr. Mark Daly, R01-MH089175 awarded to Dr. Richard Gibbs, R01-MH089025 awarded to Joseph Buxbaum, R01-MH089004 awarded to Gerard Schellenberg, and R01-MH089482 awarded to James Sutcliffe.

***phs000424:*** The Genotype-Tissue Expression (GTEx) Project was supported by the Common Fund of the Office of the Director of the National Institutes of Health (commonfund.nih.gov/GTEx). Additional funds were provided by the NCI, NHGRI, NHLBI, NIDA, NIMH, and NINDS. Donors were enrolled at Biospecimen Source Sites funded by NCI\Leidos Biomedical Research, Inc. subcontracts to the National Disease Research Interchange (10XS170), Roswell Park Cancer Institute (10XS171), and Science Care, Inc. (X10S172). The Laboratory, Data Analysis, and Coordinating Center (LDACC) was funded through a contract (HHSN268201000029C) to The Broad Institute, Inc. Biorepository operations were funded through a Leidos Biomedical Research, Inc. subcontract to Van Andel Research Institute (10ST1035). Additional data repository and project management were provided by Leidos Biomedical Research, Inc.(HHSN261200800001E). The Brain Bank was supported supplements to University of Miami grant DA006227. Statistical Methods development grants were made to the University of Geneva (MH090941 & MH101814), the University of Chicago (MH090951,MH090937, MH101825, & MH101820), the University of North Carolina -Chapel Hill (MH090936), North Carolina State University (MH101819),Harvard University (MH090948), Stanford University (MH101782), Washington University (MH101810), and to the University of Pennsylvania (MH101822).

***phs000654:*** The sequencing was performed through Epi4K Gene Discovery in Epilepsy study (NINDS U01-NS077303) and the Epilepsy Genome/Phenome Project (EPGP -NINDS U01-NS053998).

***phs000687:*** The Bulgarian Trio Sequencing study is an accumulation of exome sequencing performed and/or funded by the Broad Institute, Cardiff University, Icahn School of Medicine at Mount Sinai, and the Wellcome Trust Sanger Institute. Work at the Broad Institute was funded by Fidelity Foundations, the Sylvan Herman Foundation and philanthropic gifts from Kent and Liz Dauten, Ted and Vada Stanley, and an anonymous donor to the Stanley Center for Psychiatric Research. Work at Cardiff was supported by Medical Research Council (MRC) Centre (G0800509) and Program Grants (G0801418), the European Community’s Seventh Framework Programme (HEALTH-F2-2010-241909 (Project EU-GEI)). Work at the Icahn School of Medicine at Mount Sinai was supported by the Friedman Brain Institute, the Institute for Genomics and Multiscale Biology and National Institutes of Health grants R01HG005827 (SMP) and R01MH071681 (PS). Work at the Wellcome Trust Sanger Institute was supported by The Wellcome Trust (WT089062 and WT098051). The recruitment of the trios in Bulgaria was funded by the Janssen Research Foundation.

***phs000876:*** This research was supported in part by NCI grants U19CA148127 and P30CA023108, as well as Canadian Cancer Society Research Institute (no. 020214) and the International Agency for Research on Cancer.

***phs000971:*** This publication includes data from the ClinSeq™ study that was supported by the National Human Genome Research Institute Intramural Research Program.

***phs000806, phs001000 and phs001101:*** We thank the Broad Institute for generating high-quality sequence data supported by NHGRI funds (grant # U54 HG003067) with Eric Lander as PI.

## Author contributions

R.J.K. and Z.H.G conceived and designed the study. ZHG supervised the research. M.E and R.J.K performed data analysis. M.E, R.J.K and Z.H.G wrote and reviewed the manuscript with critical feedback and support from K.O and S.G. All authors approved the manuscript. We would like to thank the ISMMS BioMe team for providing whole-exome sequencing calls, principal component analysis, and identification of unrelated individuals.

## List of Supplementary Tables

**Supplementary Table S1.** dbGaP control cohort

**Supplementary Table S2.** Distribution of samples in TCGA

**Supplementary Table S3.** List of ICD9/10 codes for identifying cancer patients. Data in Supp_Table_S3_ICD9_10.xlsx

**Supplementary Table S4.** List of genes used in gene-set level burden analyses

**Supplementary Table S5.** Distribution of males and females in the study cohorts

**Supplementary Table S6.** List of variants identified during joint calling in discovery cohort after sample and variant QC (6,371 cases and 6,647 controls)

**Supplementary Table S7.** List of genes with significant burden in pan-cancer and tissue-specific gene level burden analysis. Data in Supp_Table_S7_genelevel.xlsx

**Supplementary Table S8.** The gene-set level burden (odds ratios (OR) and p-values)) of rare, deleterious variant in Fanconi Anemia gene-set for pan-cancer and tissue-specific cancer. Data in Supp_Table_S8_FAgenesetlevel.xlsx

**Supplementary Table S9.** Gene-set level rare, deleterious variant (RDV) burden in the study cohorts without BRCA1/2 RDVs

**Supplementary Table S10.** The gene-set level burden (odds ratios (OR) and p-values) of rare, deleterious variant for all gene-sets listed in Molecular Signatures Database (MSigDB) for pan-cancer. Data in Supp_Table_S10_MSigDB.xlsx

**Supplementary Table S11.** Comparison of immune cell fractions (from CIBERSORT) based on germline RDV load in TCGA cases for different gene-sets. Data in Supp_Table_S11_TME.xlsx

**Supplementary Table S12.** Comparison of gene expression of genes related to M1 Macrophages (LM22 matrix) based on germline RDV load in TCGA cases for different gene-sets. Data in Supp_Table_S12_M1Macrophages_Genes.xlsx

**Supplementary Table S13.** Effect of M1 Macrophages on survival in TCGA cases. Data in Supp_Table_S13_Survival.xlsx

**Supplementary Table S14.** Comparison of tumor mutational burden (TMB) based on germline RDV load in TCGA cases for different gene-sets. Data in Supp_Table_S14_TMB.xlsx

## List of Supplementary Figures

**Supplementary Fig. S1.** Gating of individuals of European ancestry in the BioMe Cohort

**Supplementary Fig. S2.** Tally of genes with per-sample rare synonymous variants between cases and controls

**Supplementary Fig. S3.** Effect of gender on gene-set level burden of rare, deleterious variant in the study cohorts

**Supplementary Fig. S4.** Comparison of immune cell fractions (from CIBERSORT) based on germline RDV load in TCGA cases for different gene-sets

