## Supplemental File for "Germline rare deleterious variant load alters cancer risk, age of onset and tumor characteristics"

**List of Supplementary Tables**
**Supplementary Table S1.** dbGaP control cohort

**Supplementary Table S2.** Distribution of samples in TCGA

**Supplementary Table S3.** List of ICD9/10 codes for identifying cancer patients. Data in Supp_Table_S3_ICD9_10.xlsx

**Supplementary Table S4.** List of genes used in gene-set level burden analyses

**Supplementary Table S5.** Distribution of males and females in the study cohorts

**Supplementary Table S6.** List of variants identified during joint calling in discovery cohort after sample and variant QC (6,371 cases and 6,647 controls)

**Supplementary Table S7.** List of genes with significant burden in pan-cancer and tissue-specific gene level burden analysis. Data in Supp_Table_S7_genelevel.xlsx

**Supplementary Table S8.** The gene-set level burden (odds ratios (OR) and p-values)) of rare, deleterious variant in Fanconi Anemia gene-set for pan-cancer and tissue-specific cancer. Data in Supp_Table_S8_FAgenesetlevel.xlsx

**Supplementary Table S9.** Gene-set level rare, deleterious variant (RDV) burden in the study cohorts without BRCA1/2 RDVs

**Supplementary Table S10.** The gene-set level burden (odds ratios (OR) and p-values) of rare, deleterious variant for all gene-sets listed in Molecular Signatures Database (MSigDB) for pan-cancer. Data in Supp_Table_S10_MSigDB.xlsx

**Supplementary Table S11.** Comparison of immune cell fractions (from CIBERSORT) based on germline RDV load in TCGA cases for different gene-sets. Data in Supp_Table_S11_TME.xlsx

**Supplementary Table S12.** Comparison of gene expression of genes related to M1 Macrophages (LM22 matrix) based on germline RDV load in TCGA cases for different gene-sets. Data in Supp_Table_S12_M1Macrophages_Genes.xlsx

**Supplementary Table S13.** Effect of M1 Macrophages on survival in TCGA cases. Data in Supp_Table_S13_Survival.xlsx

**Supplementary Table S14.** Comparison of tumor mutational burden (TMB) based on germline RDV load in TCGA cases for different gene-sets. Data in Supp_Table_S14_TMB.xlsx

**List of Supplementary Figures**

**Supplementary Fig. S1.** Gating of individuals of European ancestry in the BioMe Cohort

**Supplementary Fig. S2.** Tally of genes with per-sample rare synonymous variants between cases and controls

**Supplementary Fig. S3.** Effect of gender on gene-set level burden of rare, deleterious variant burden in the study cohorts

**Supplementary Fig. S4.** Comparison of immune cell fractions (from CIBERSORT) based on germline RDV load in TCGA cases for different gene-sets

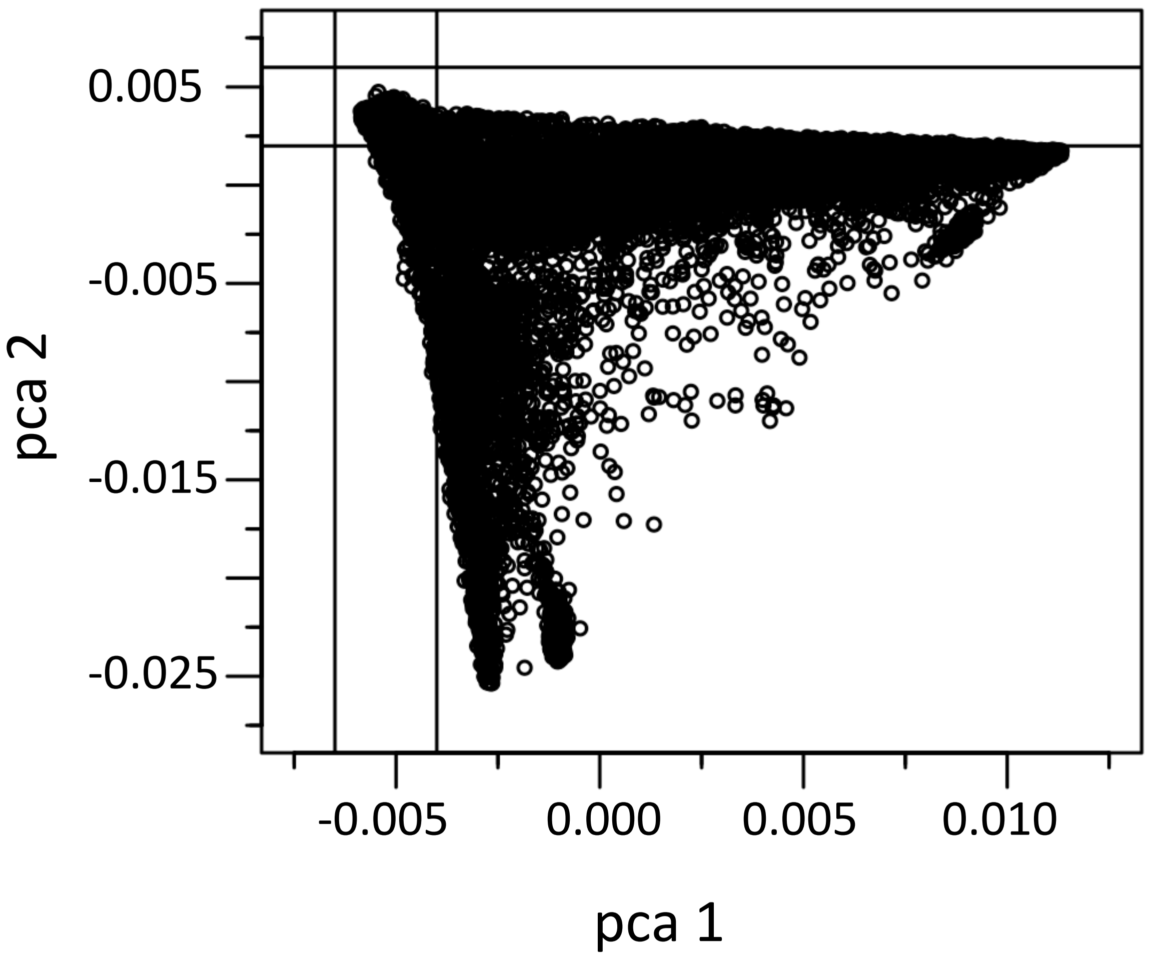


**Figure S1. Gating individuals of European ancestry in the BioMe Cohort.** 10,784 individuals of European ancestry were gated using the top two principal components.

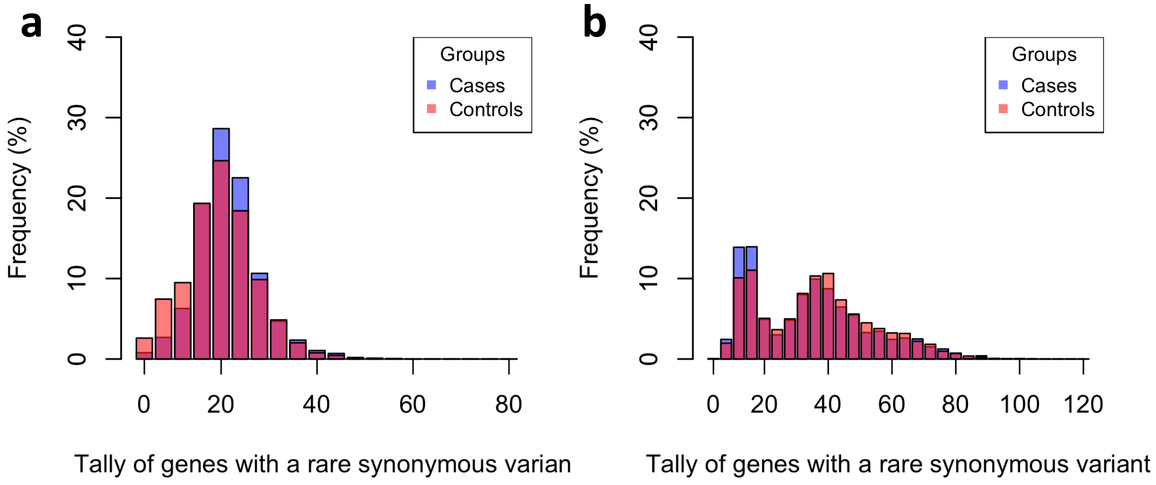


**Figure S2: Tally of genes with per-sample rare synonymous variants between cases and controls**. **A)** Discovery cohort, Cases: average 19.98±6.7 genes, Controls: average 18.38±7.3 genes, Mann-Whitney U test p-value < 2.2e-16. **B)** Validation cohort, Cases: average 19.98±6.7 genes, Controls: average 18.38±7.3 genes, Mann-Whitney U test p-value < 2.2e-16.

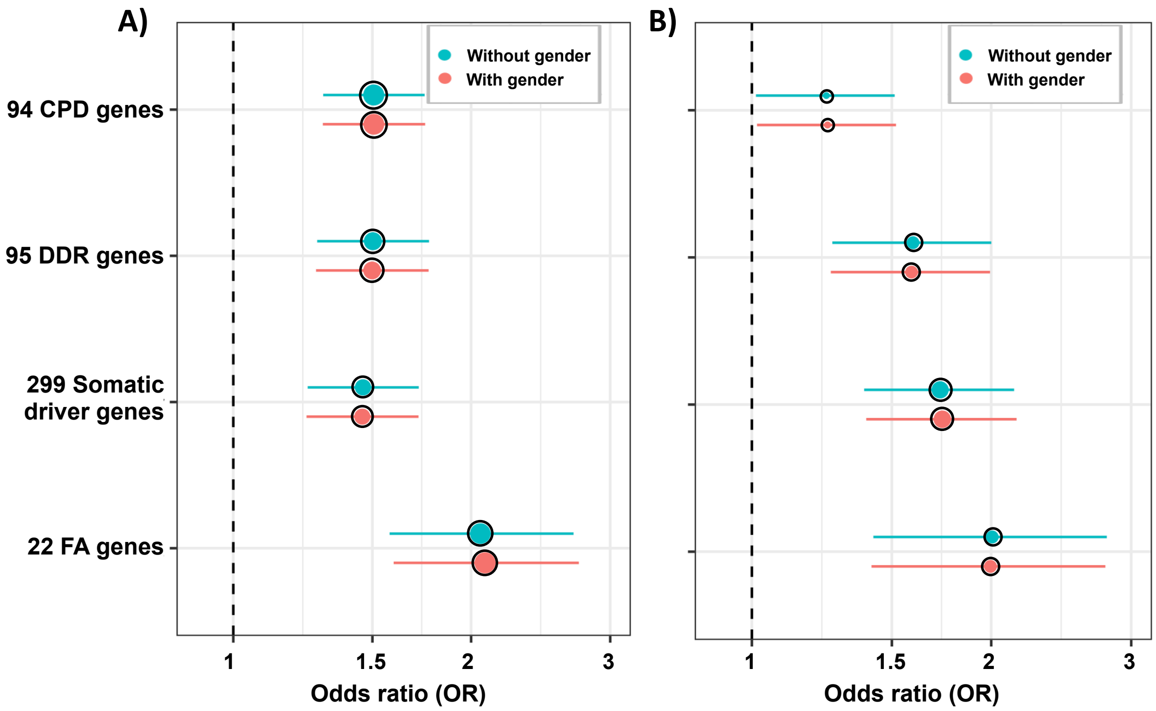


**Figure S3: Effect of gender on gene-set level burden of rare, deleterious variant in the study cohorts.** Gene-set level burden of rare, deleterious variants when using Penalized Logistic Regression Analysis (PLRA) (i) without gender effect, by not using gender information as a covariate (blue) and (ii) with gender effect, by using the gender information as additional covariate (red). **A)** Discovery cohort; **B)** Validation cohort. The whiskers span the 95% confidence interval for OR values. The black circle outline indicates significant burden p ≤ 0.05

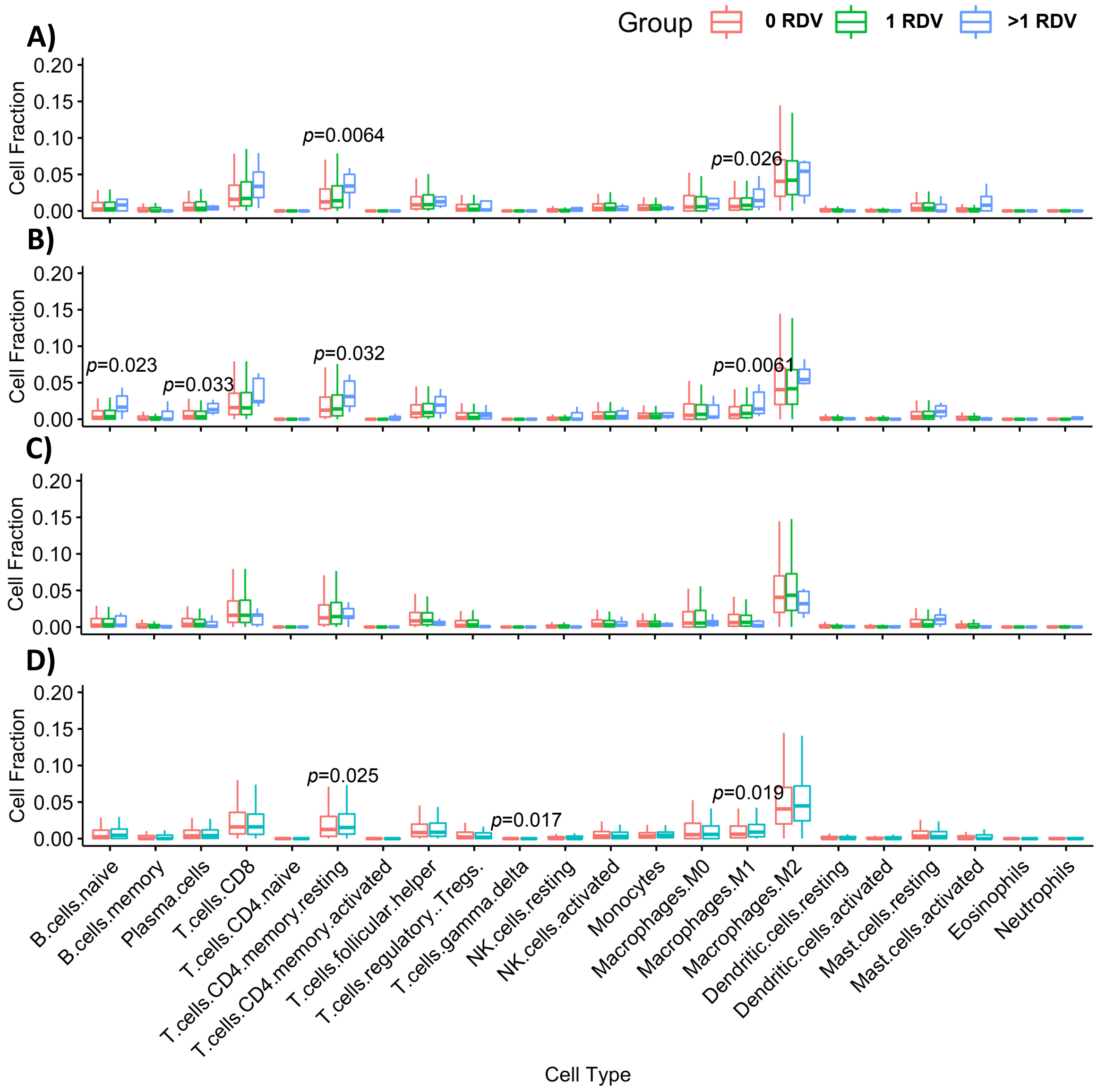


**Figure S4. Comparison of immune cell fractions (from CIBERSORT) based on germline RDV load in TCGA cases for different gene-sets. A)** 94 Cancer predisposition genes **B)** 95 DNA damage repair genes **C)** 299 Somatic cancer driver genes **D)** 22 Fanconi Anemia genes. Significant *p* values from Kruskal-Wallis test are included in the figure.

**Table S1: dbGaP control cohort**. Total number of samples in each dbGaP study before and after sample QC.

| dbGaP Study Accession | Study Name | Start Count | Final Count |
|---|---|---|---|
| phs000209 | Multi-Ethnic Study of Atherosclerosis (MESA) Cohort | 401 | 242 |
| phs000276 | STAMPEED: Northern Finland Birth Cohort 1966 (NFBC1966) | 514 | 513 |
| phs000296 | NHLBI GO-ESP: Lung Cohorts Exome Sequencing Project (COPDGene) | 290 | 280 |
| phs000298 | ARRA Autism Sequencing Collaboration | 168 | 168 |
| phs000424 | Common Fund (CF) Genotype-Tissue Expression Project (GTEx) | 478 | 380 |
| phs000654 | Genetic Analyses in Epileptic Encephalopathies | 583 | 457 |
| phs000687 | Bulgarian Trio Sequencing Study to Identify de Novo Mutations in Schizophrenia | 555 | 524 |
| phs000806 | Myocardial Infarction Genetics Exome Sequencing Consortium: Ottawa Heart Study | 964 | 942 |
| phs000876 | Transdisciplinary Research Into Cancer of the Lung (TRICL) - Exome Plus Targeted Sequencing | 591 | 551 |
| phs000971 | The ClinSeq Project: Piloting large-scale genome sequencing for research in genomic medicine | 621 | 519 |
| phs001000 | Myocardial Infarction Genetics Exome Sequencing Consortium: U. of Leicester | 1153 | 1022 |
| phs001101 | Myocardial Infarction Genetics Exome Sequencing Consortium: Malmo Diet and Cancer Study | 1070 | 1049 |
| **Total** | | 7388 | 6647 |

**Table S2: Distribution of samples in TCGA**

| Sl.No | Cancer Type | # Cases before QC | # Cases after QC |
|---|---|---|---|
| 1 | Breast invasive carcinoma | 848 | 622 |
| 2 | Lung adenocarcinoma | 546 | 463 |
| 3 | Lung squamous cell carcinoma | 494 | 443 |
| 4 | Brain Lower Grade Glioma | 508 | 426 |
| 5 | Head and Neck squamous cell carcinoma | 526 | 426 |
| 6 | Skin Cutaneous Melanoma | 453 | 424 |
| 7 | Prostate adenocarcinoma | 491 | 403 |
| 8 | Thyroid carcinoma | 487 | 338 |
| 9 | Uterine Corpus Endometrial Carcinoma | 480 | 331 |
| 10 | Bladder Urothelial Carcinoma | 400 | 305 |
| 11 | Kidney renal clear cell carcinoma | 367 | 279 |
| 12 | Stomach adenocarcinoma | 415 | 268 |
| 13 | Colon adenocarcinoma | 350 | 267 |
| 14 | Glioblastoma multiforme | 310 | 252 |
| 15 | Ovarian serous cystadenocarcinoma | 277 | 235 |
| 16 | Kidney renal papillary cell carcinoma | 280 | 193 |
| 17 | Liver hepatocellular carcinoma | 363 | 167 |
| 18 | Cervical SqCC and endocervical adenocarcinoma | 283 | 162 |
| 19 | Rectum adenocarcinoma | 107 | 99 |
| 20 | Adrenocortical carcinoma | 90 | 76 |
| 21 | Acute Myeloid Leukemia | 75 | 67 |
| 22 | Kidney Chromophobe | 66 | 56 |
| 23 | Uterine Carcinosarcoma | 57 | 42 |
| 24 | Lymphoid Neoplasm Diffuse Large B-cell Lymphoma | 48 | 27 |
| | ***Total*** | 8321 | 6371 |

**Table S4. List of genes used in gene-set level burden analyses**.

| | Gene Symbol |
|---|---|
| ***Gene Set I:*** ***94 TruSight risk panel genes*** | *AIP, ALK, APC, ATM, BAP1, BLM, BMPR1A, BRCA1, BRCA2, BRIP1, BUB1B, CDC73, CDH1, CDK4, CDKN1C, CDKN2A, CEBPA, CEP57, CHEK2, CYLD, DDB2, DICER1, DIS3L2, EGFR, EPCAM, ERCC2, ERCC3, ERCC4, BIVM-ERCC5, EXT1, EXT2, EZH2, FANCA, FANCB, FANCC, FANCD2, FANCE, FANCF, FANCG, FANCI, FANCL, FANCM, FH, FLCN, GATA2, GPC3, HNF1A, HRAS, KIT, MAX, MEN1, MET, MLH1, MSH2, MSH6, MUTYH, NBN, NF1, NF2, NSD1, PALB2, PHOX2B, PMS1, PMS2, PRF1, PRKAR1A, PTCH1, PTEN, RAD51C, RAD51D, RB1, RECQL4, RET, RHBDF2, RUNX1, SBDS, SDHAF2, SDHB, SDHC, SDHD, SLX4, SMAD4, SMARCB1, STK11, SUFU, TMEM127, TP53, TSC1, TSC2, VHL, WRN, WT1, XPA, XPC* |
| ***Gene Set II:*** ***95 DNA repair genes*** | *ALKBH2, ALKBH3, APEX1, APEX2, ATM, ATR, ATRIP, BAP1, BARD1, BIVM-ERCC5, BLM, BRCA1 (FANCS), BRCA2 (FANCD1), BRIP1 (FANCJ), CHEK1, CHEK2, CUL5, EME1, ERCC1, ERCC2, ERCC4 (FANCQ), ERCC6, EXO1, FAM175A, FANCA, FANCB, FANCC, FANCD2, FANCE, FANCF, FANCG, FANCI, FANCL, FANCM, FEN1, GEN1, LIG4, MAD2L2, MDC1, MGMT, MLH1, MLH3, MRE11A, MSH2, MSH3, MSH6, MUS81, NBN, NHEJ1, NUDT1, NUDT15, NUDT18, PALB2 (FANCN), PARP1, PMS1, PMS2, POLB, POLE, POLE3, POLL, POLM, POLN, POLQ, PRKDC, RAD50, RAD51 (FANCR), RAD51C (FANCO), RAD51D, RAD52, RBBP8, REV1, REV3L, RFWD3 (FANCW), RNMT, RRM1, RRM2, SHFM1, SHPRH, SLX1A, SLX4 (FANCP), TDG, TDP1, TOP3A, TOPBP1, TP53BP1, TREX1, UBE2T (FANCT), UNG, XPA, XPC, XRCC2 (FANCU), XRCC3, XRCC4, XRCC5, XRCC6* |
| ***Gene Set III:*** ***299 cancer driver genes*** | *ABL1, ACVR1, ACVR1B, ACVR2A, AJUBA, AKT1, ALB, ALK, AMER1, APC, APOB, AR, ARAF, ARHGAP35, ARID1A, ARID2, ARID5B, ASXL1, ASXL2, ATF7IP, ATM, ATR, ATRX, ATXN3, AXIN1, AXIN2, B2M, BAP1, BCL2, BCL2L11, BCOR, BRAF, BRCA1, BRCA2, BRD7, BTG2, CACNA1A, CARD11, CASP8, CBFB, CBWD3, CCND1, CD70, CD79B, CDH1, CDK12, CDK4, CDKN1A, CDKN1B, CDKN2A, CDKN2C, CEBPA, CHD3, CHD4, CHD8, CHEK2, CIC, CNBD1, COL5A1, CREB3L3, CREBBP, CSDE1, CTCF, CTNNB1, CTNND1, CUL1, CUL3, CYLD, CYSLTR2, DACH1, DAZAP1, DDX3X, DHX9, DIAPH2, DICER1, DMD, DNMT3A, EEF1A1, EEF2, EGFR, EGR3, EIF1AX, ELF3, EP300, EPAS1, EPHA2, EPHA3, ERBB2, ERBB3, ERBB4, ERCC2, ESR1, EZH2, FAM46D, FAT1, FBXW7, FGFR1, FGFR2, FGFR3, FLNA, FLT3, FOXA1, FOXA2, FOXQ1, FUBP1, GABRA6, GATA3, GNA11, GNA13, GNAQ, GNAS, GPS2, GRIN2D, GTF2I, H3F3A, H3F3C, HGF, HIST1H1C, HIST1H1E, HLA-A, HLA-B, HRAS, HUWE1, IDH1, IDH2, IL6ST, IL7R, INPPL1, IRF2, IRF6, JAK1, JAK2, JAK3, KANSL1, KDM5C, KDM6A, KEAP1, KEL, KIF1A, KIT, KLF5, KMT2A, KMT2B, KMT2C, KMT2D, KRAS, KRT222, LATS1, LATS2, LEMD2, LZTR1, MACF1, MAP2K1, MAP2K4, MAP3K1, MAP3K4, MAPK1, MAX, MECOM, MED12, MEN1, MET, MGA, MGMT, MLH1, MSH2, MSH3, MSH6, MTOR, MUC6, MYC, MYCN, MYD88,* |

| | |
|---|---|
| | *MYH9, NCOR1, NF1, NF2, NFE2L2, NIPBL, NOTCH1, NOTCH2, NPM1, NRAS, NSD1, NUP133, NUP93, PAX5, PBRM1, PCBP1, PDGFRA, PDS5B, PGR, PHF6, PIK3CA, PIK3CB, PIK3CG, PIK3R1, PIK3R2, PIM1, PLCB4, PLCG1, PLXNB2, PMS1, PMS2, POLE, POLQ, POLRMT, PPM1D, PPP2R1A, PPP6C, PRKAR1A, PSIP1, PTCH1, PTEN, PTMA, PTPDC1, PTPN11, PTPRC, PTPRD, RAC1, RAD21, RAF1, RARA, RASA1, RB1, RBM10,, RET, RFC1, RHEB, RHOA, RHOB, RIT1, RNF111, RNF43, RPL22, RPL5, RPS6KA3, RQCD1, RRAS2, RUNX1, RXRA, SCAF4, SETBP1, SETD2, SF1, SF3B1, SIN3A, SMAD2, SMAD4, SMARCA1, SMARCA4, SMARCB1, SMC1A, SMC3, SOS1, SOX17, SOX9, SPOP, SPTA1, SPTAN1, SRSF2, STAG2, STK11, TAF1, TBL1XR1, TBX3, TCEB1, TCF12, TCF7L2, TET2, TGFBR2, TGIF1, THRAP3, TLR4, TMSB4X, TNFAIP3, TP53, TRAF3, TSC1, TSC2, TXNIP, U2AF1, UNCX, USP9X, VHL, WHSC1, WT1, XPO1, ZBTB20, ZBTB7B, ZC3H12A, ZCCHC12, ZFHX3, ZFP36L1, ZFP36L2, ZMYM2, ZMYM3, ZNF133, ZNF750* |
| ***Gene Set IV:***<br>***22 Fanconi Anemia genes*** | *FANCA, FANCB, FANCC, FANCD1 (BRCA2), FANCD2, FANCE, FANCF, FANCG, FANCI, FANCJ (BRIP1), FANCL, FANCM, FANCN (PALB2), FANCO (RAD51C), FANCP (SLX4), FANCQ (ERCC4), FANCR (RAD51), FANCS (BRCA1), FANCT (UBE2T), FANCU (XRCC2), FANCV (REV7), FANCW (RFWD3)* |

**Table S5: Distribution of males and females in the study cohorts.**

**A). Discovery cohort**

| Sl.No | Cancer Type | Sample | Male | Female | Missing |
|---|---|---|---|---|---|
| 1 | Breast invasive carcinoma | 622 | 6 (0.96%) | 616 (99.04%) | 0 |
| 2 | Lung adenocarcinoma | 463 | 196 (42.33%) | 231 (49.89%) | 36 (7.78%) |
| 3 | Lung squamous cell carcinoma | 443 | 333 (75.17%) | 110 (24.83%) | 0 |
| 4 | Brain Lower Grade Glioma | 426 | 237 (55.63%) | 188 (44.13%) | 1 (0.23%) |
| 5 | Head and Neck squamous cell carcinoma | 426 | 305 (71.60%) | 121 (28.40%) | 0 |
| 6 | Skin Cutaneous Melanoma | 424 | 258 (60.85%) | 166 (39.15%) | 0 |
| 7 | Prostate adenocarcinoma | 403 | 403 (100%) | 0 | 0 |
| 8 | Thyroid carcinoma | 338 | 92 (27.22%) | 246 (72.78%) | 0 |
| 9 | Uterine Corpus Endometrial Carcinoma | 331 | 0 | 328 (99.09%) | 3 (0.91%) |
| 10 | Bladder Urothelial Carcinoma | 305 | 228 (74.75%) | 77 (25.25%) | 0 |
| 11 | Kidney renal clear cell carcinoma | 279 | 183 (65.59%) | 96 (34.41%) | 0 |
| 12 | Stomach adenocarcinoma | 268 | 159 (59.33%) | 109 (40.67%) | 0 |
| 13 | Colon adenocarcinoma | 267 | 144 (53.93%) | 123 (46.07%) | 0 |
| 14 | Glioblastoma multiforme | 252 | 159 (63.10%) | 92 (36.51%) | 1 (0.40%) |
| 15 | Ovarian serous cystadenocarcinoma | 235 | 0 | 235 (100%) | 0 |
| 16 | Kidney renal papillary cell carcinoma | 193 | 146 (75.65%) | 47 (24.35%) | 0 |
| 17 | Liver hepatocellular carcinoma | 167 | 92 (55.09%) | 75 (44.91%) | 0 |
| 18 | Cervical SqCC and endocervical adenocarcinoma | 162 | 0 | 162 (100%) | 0 |
| 19 | Rectum adenocarcinoma | 99 | 52 (52.53%) | 46 (46.46%) | 1 (1.01%) |
| 20 | Adrenocortical carcinoma | 76 | 28 (36.84%) | 48 (63.16%) | 0 |
| 21 | Acute Myeloid Leukemia | 67 | 34 (50.75%) | 33 (49.25%) | 0 |
| 22 | Kidney Chromophobe | 56 | 33 (58.93%) | 23 (41.07%) | 0 |
| 23 | Uterine Carcinosarcoma | 42 | 0 | 42 (100%) | 0 |
| 24 | Lymphoid Neoplasm Diffuse Large B-cell Lymphoma | 27 | 11 (40.74%) | 16 (59.26%) | 0 |
| | **Total cases** | 6371 | 3099 (48.64%) | 3230 (50.70%) | 42 (0.66%) |
| | **Total controls** | 6647 | 4034 | 2610 | 3 |

| | | | | | |
|---|---|---|---|---|---|
| | | | (60.69%) | (39.27%) | (0.05%) |

**B). Validation cohort**

| | Male | Female |
|---|---|---|
| Cases (1,571) | 690 (43.92%) | 881 (56.08%) |
| Controls (6,200) | 3150 (50.81%) | 3050 (49.19%) |

**Table S6. List of variants identified during joint calling in discovery cohort after sample and variant QC (6,371 cases and 6,647 controls)**

| | Variant Type | Number of sites |
|---|---|---|
| 1 | Exonic | 935,912 |
| 2 | Exonic; splicing | 245 |
| 3 | ncRNA_exonic; splicing | 18 |
| 4 | ncRNA_splicing | 21 |
| 5 | Splicing | 5,413 |
| | | 941,609 |

**Table S9. Gene-set level rare, deleterious variant (RDV) burden in the study cohorts without BRCA1/2 RDVs**

| | TCGA-dbGaP Cohort | | BioMe Cohort | |
|---|---|---|---|---|
| | **Cases (6,371)** | **Controls (6,647)** | **Cases (1,571)** | **Controls (6,200)** |
| | **92 CPD genes** | | | |
| # Variants | 226 | 168 | 67 | 162 |
| # Genes | 55 | 46 | 33 | 49 |
| # Unique individuals | 392 (6.15%) | 300 (4.51%) | 101 (6.43%) | 384 (6.19%) |
| OR (*p*-value) (95% CI) | 1.37 (7.61e-05) [1.17-1.60] | | 1.05 (0.68) [0.83-1.31] | |
| | **93 DDR genes** | | | |
| # Variants | 206 | 163 | 61 | 130 |
| # Genes | 39 | 34 | 27 | 33 |
| # Unique individuals | 300 (4.71%) | 243 (3.66%) | 71 (4.52%) | 217 (3.50%) |
| OR (*p*-value) (95% CI) | 1.33 (1.54e-03) [1.11-1.58] | | 1.32 (0.05) [1.00-1.72] | |
| | **297 Somatic Driver Genes** | | | |
| # Variants | 183 | 136 | 67 | 139 |
| # Genes | 57 | 50 | 38 | 57 |
| # Unique individuals | 307 (4.82%) | 247 (3.72%) | 85 (5.41%) | 236 (3.81%) |
| OR (*p*-value) (95% CI) | 1.30 (3.3e-03) [1.09-1.54] | | 1.45 (4.94e-03) [1.12-1.87] | |
| | **20 Fanconi Anemia Genes** | | | |
| # Variants | 67 | 40 | 11 | 39 |
| # Genes | 13 | 11 | 6 | 11 |
| # Unique individuals | 88 (1.38%) | 59 (0.89%) | 12 (0.76%) | 46 (0.74%) |
| OR (*p*-value) (95% CI) | 1.60 (5.41e-03) [1.15-2.25] | | 1.11 (0.75) [0.57-2.01] | |
